# Prospective study of machine learning for identification of high-risk COVID-19 patients

**DOI:** 10.1101/2024.02.21.24303159

**Authors:** Mateo Frausto-Avila, Roberto de J. León-Montiel, Mario A. Quiroz-Juárez, Alfred B. U’Ren

**Affiliations:** Centro de Física Aplicada y Tecnología Avanzada, Universidad Nacional Autónoma de México, Boulevard Juriquilla 3001, 76230 Querétaro, México; Instituto de Ciencias Nucleares, Universidad Nacional Autónoma de México, Apartado Postal 70-543, 04510 Cd. Mx., México

## Abstract

The Coronavirus Disease 2019 (COVID-19) pandemic constituted a public health crisis with a devastating effect in terms of its death toll and effects on the world economy. Notably, machine learning methods have played a pivotal role in devising novel technological solutions designed to tackle challenges brought forth by this pandemic. In particular, tools for the rapid identification of high-risk COVID-19 patients have been developed to aid in the effective allocation of hospital resources and for containing the spread of the virus. A comprehensive validation of such intelligent technological approaches is needed to ascertain their clinical utility; importantly, it may help develop future strategies for efficient patient classification to be used in future viral outbreaks. Here we present a prospective study to evaluate the performance of state-of-the-art machine-learning models proposed in PloS one 16, e0257234 (2021), which we developed for the identification of high-risk COVID-19 patients across four identified clinical stages. The model relies on artificial neural networks trained with historical patient data from Mexico. To assess their predictive capabilities across the six, registered, epidemiological waves of COVID-19 infection in Mexico, we measure the accuracy within each wave without retraining the neural networks. We then compare their performance against neural networks trained with cumulative historical data up to the end of each wave. Our findings indicate that models trained using early historical data exhibit strong predictive capabilities, which allows us to accurately identify high-risk patients in subsequent epidemiological waves—under clearly varying vaccination, prevalent viral strain, and medical treatment conditions. These results show that artificial intelligence-based methods for patient classification can be robust throughout an extended period characterized by constantly evolving conditions, and represent a potentially powerful tool for tackling future pandemic events, particularly for clinical outcome prediction of individual patients.

## I. Introduction

A report of several cases of viral pneumonia by the Wuhan Municipal Health Commission in China on 12 December 2019 evolved into the declaration of the COVID-19 pandemic by the World Health Organization on 11 March 2020 [1, 2]. The profound impact of COVID-19 on a global scale is attributed to its highly contagious nature and substantial mortality rate. This infectious disease has not only inflicted a severe toll on the global population but has also imposed immense challenges on governments and the world economy [3, 4]. The ensuing strain has compelled nations to navigate unprecedented difficulties in their efforts to contain the virus and mitigate its far-reaching consequences. The crisis has underscored the inadequacies of healthcare systems globally, revealing critical deficiencies in hospital equipment, medical personnel, and overall healthcare infrastructure. This has served as a testament to the systemic vulnerabilities and the urgent need for comprehensive emergency preparedness and response strategies on a global scale [5–7].

Several different strategies have been explored to tackle challenges associated with logistics, supply chain management, and the mathematical modeling of viral spread [8–16]. These endeavors have been aimed at preemptively addressing or alleviating deficiencies in emergency-response systems and healthcare infrastructures. In particular, the identification of high-risk patients is important because hospital resources and capacities must be adequately managed to prevent the collapse of healthcare systems [17]. In this direction, several approaches based on machine-learning algorithms have been proposed to identify, from the earliest stage possible, patients who are likely to become ill or critically ill. These approaches make predictions relying on basic patient information [18–23], clinical symptoms [24, 25], as well as travel history [26] and the discharge time of hospitalized patients [27]. Some other efforts focus on identifying patients that require specialized care, namely hospitalization and/or special care units [28, 29], or patients at a higher fatality risk [30].

Further studies have been reported that investigate the dynamics of epidemiological waves, thus addressing the complexity of viral spread [31, 32]. Despite notable advances in the development of machine learning-based algorithms for the early identification of high-risk patients [33], it is crucial to highlight the importance of conducting prospective studies that validate the effectiveness and accuracy of these algorithms in real-world situations. The application of prospective approaches will thus significantly contribute to the evaluation of the clinical utility of such tools, thus ensuring their reliability in practical settings for the improvement of medical decision-making during health emergencies. The successful implementation of such tools may improve resource management in hospitals and healthcare units in the event of future health crises.

In this work, we present a prospective validation study of the state-of-the-art machine-learning models that we proposed in Ref. [17], designed for the identification of high-risk COVID-19 patients, across four clinical stages, in Mexico. Our study is based on a patient database, publicly made available by the Mexican federal government, covering the period from the 10th week of 2020 to the 13th week of 2023, which includes i) demographic, ii) COVID-19 status, and iii) comorbidity information for patients known or suspected to have been infected with COVID-19, as reported from within the Mexican healthcare system. Importantly, the treatment outcome (i.e. recovery or death) is available for each patient on this database. To evaluate the predictive performance of our models through the six COVID-19 epidemiological waves identified in Mexico, we determine the algorithm accuracy for patients within each wave without any retraining of the neural networks reported in Ref. [17]. Subsequently, we compare their effectiveness against neural networks trained with cumulative historical data, covering the period up to the end of each wave. Our results indicate that models trained with early historical data exhibit strong predictive capabilities throughout all subsequent epidemiological waves. This demonstrates that artificial intelligence algorithms can not only provide accurate identification of high-risk patients based on limited data, but may be robust despite a constantly evolving set of conditions, particularly in terms of the population vaccination status, the dominant viral strains, and the available medical treatments. This paper is structured as follows: Section II describes the database used for this study, Section III presents a detailed description of our findings, and Section IV is devoted to our conclusions.

## II. Materials and methods

### A. Data

The prospective validation study was conducted using the publicly available database of COVID-19 patients in Mexico. This database, which includes all officially reported confirmed and suspected cases of COVID-19 in Mexico, is available in the Statistical Yearbook of Disease (Anuario Estadísticos de Morbilidad) published by the General Epidemiological Council (Dirección General de Epidemiología) that is part of the Ministry of Health (Secretaría de Salud) of the Federal Government of Mexico [34].

As described in Ref. [17], each patient profile in the database comprises 28 characteristics. However, for the sake of effectiveness and accuracy, only those characteristics demonstrating sufficient predictive power were included in our analysis. Consequently, in our analysis the resulting input vector for neural networks comprised 21 features, categorized into three groups: 1) medical history, 2) demographic data, and 3) recent medical information. Category 1 includes diabetes, chronic obstructive pulmonary disease (COPD), use of immunosuppressive drugs, hypertension, chronic renal failure, and cardiovascular diseases. Category 2 includes gender, age, state of birth, state of residence, and age. Category 3 comprises health monitoring units of viral respiratory disease (USMER) designation, type of health-care facility where the patient is receiving treatment, state of treatment, number of days between the onset of symptoms and the beginning of treatment, COVID-19 status, COVID-19-related pneumonia, hospitalization status, intubation and admission into an intensive care unit (ICU).

In this prospective study, we conduct patient tracking through each epidemiological wave to assess the predictive capabilities of the models proposed by Ref. [17]. These models were trained using data covering the period up to 31 January 2021. As of the present writing, it has the following attributes:

a. **Recorded dates.-** The database covers the period from May 12 2020 to April 4 2023. In this period, it contains the historical record of 25,118,719 patients. Figure 1(a) shows an overview of the COVID-19 pandemic in Mexico. For each wave, the cumulative epidemic percentage (CEP), epidemic wave percentage (EWP), and cumulative percentage in the Mexican population (CPM) are presented.
b. **Epidemiological waves.-** During the pandemic Mexico officially experienced six epidemiological waves (EWs) with variable duration and four inter-pandemic periods (IPs). Figure 1(a) illustrates each EW in blue, while IPs are highlighted in gray. The corresponding periods and duration, expressed in weeks for both, EWs and IPs, are presented in Table I.
c. **Database sampling.-** The database contains daily record updates up to the period IP-4. Unfortunately, in the beginning of this inter-pandemic period, the sampling frequency became irregular —attributed to the low prevalence of cases—until eventually updates to the database ceased. This implies that when carrying out a prospective study for patients in EW-5, there is uncertainty as to the specific day when a patient transitions from one stage to another.
d. **Survivors and Death Tolls.-** As of the date of the last update to the database, 456,252 deaths had been recorded among the 25,118,719 patients included in the historical database. Figure 1(b) shows a cumulative histogram depicting the total number of deaths experienced within each epidemiological wave during the entire COVID-19 outbreak.

**TABLE I.** Epidemiological waves (EW) and inter-pandemic periods (IP) in Mexico from the 10th week of 2020 to the 13th week of 2023.

| Event | Period (weeks) | Duration (weeks) |
| --- | --- | --- |
| EW-1 | 10 to 39 of 2020 | 31 |
| EW-2 | 39 of 2020 to 15 of 2021 | 28 |
| IP-1 | 15 to 22 of 2021 | 07 |
| EW-3 | 22 to 42 of 2021 | 20 |
| IP-2 | 42 to 50 of 2021 | 08 |
| EW-4 | 50 of 2021 to 09 of 2022 | 11 |
| IP-3 | 09 to 21 of 2022 | 12 |
| EW-5 | 21 to 33 of 2022 | 12 |
| IP-4 | 33 to 48 of 2022 | 15 |
| EW-6 | 48 of 2022 to 13 of 2023 | 18 |

It is important to comment on the COVID-19 vaccination campaign in Mexico, which formally started on 24 december 2024. The latest vaccination record is dated 7 October 2022, indicating that by that date 76.04% of the Mexican population had received at least one vaccination dose. Thus, in the period encompassed between midway through EW-2 (when the campaign started) and the end of EW-5, slightly more than three-quarters of the population received a vaccination [35]. By inspecting this information, one can obtain specific dates corresponding to significant percentiles within the vaccinated population. The 25th, 50th, and 75th percentiles received their vaccinations on June 5, 2021, November 15, 2021, and April 27, 2022, respectively. Figure 1(a) illustrates this information in red rectangles, together with the cumulative percentage with respect to the country’s population who have received at least one vaccination dose.

**FIG. 1.**
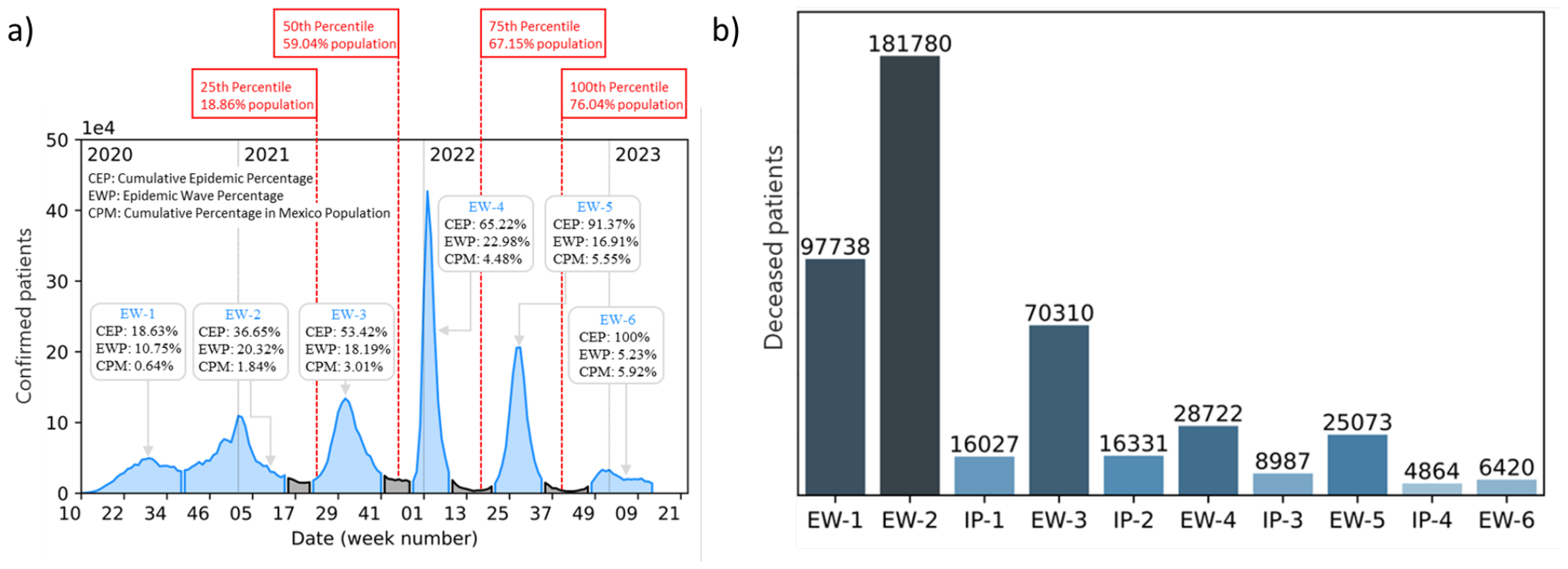
**(a)** Epidemic curve of confirmed cases in Mexico from week 10 of 2020 to week 13 of 2023, spanning a total of 162 weeks. Blue regions correspond to epidemiological waves (EW), while gray regions indicate inter-pandemic periods (IP). Additionally, vaccination records for persons who received at least one dose of complete treatment are indicated in the red rectangles. The initials used in insets correspond to CEP: cumulative epidemic percentage, EWP: epidemic wave percentage, and CPM: cumulative percentage in the Mexican population. **(b)** Recorded deaths in Mexico during the COVID-19 outbreak.

### B. Neural Network

Supervised machine learning provides computer algorithms with the ability to *learn* from a known dataset to identify features and generate predictions about the outcome given a specific set of features, not included in the learning stage. In this context, by making use of the publicly available database of Mexican COVID-19 patients, reference [17] reported on an artificial neural network capable of classifying patients into two classes: a) patients who are more likely to recover than to die or b) patients who are more likely to die than to recover. We have identified four clinical stages of the treatment process, each including a specific number of characteristics, as illustrated in the top panels of Fig. 2. Stage 1 involves patients undergoing an initial medical evaluation and/or treatment. In Stage 2, patients have a confirmed COVID-19 status as part of the medical evaluation, and may already present COVID-19-related pneumonia. Patients in stage 3 have either already been admitted to a hospital, or have returned home after a hospital stay. Patients in Stage 4 are those who have been intubated or admitted into an ICU unit. This categorization into clinical stages led to the training of four separate neural networks, one for each stage, using information dated up to 31 January 2021 from the database described in subsection IIA.

**FIG. 2.**
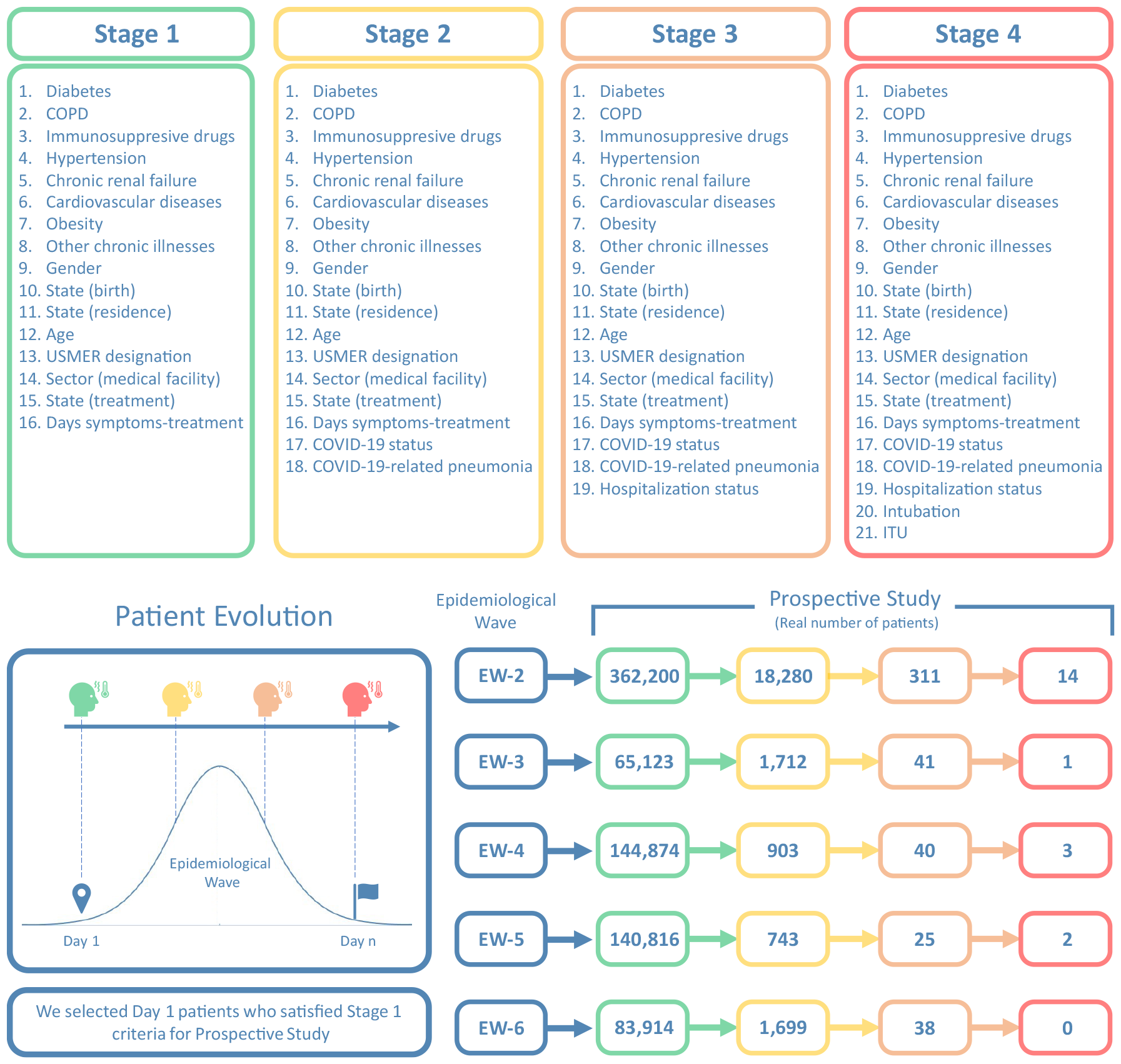
The top panels present the four clinical stages that were identified in Ref. [17], for each of which we train a separate neural network designed to identify high-risk COVID patients. The flow diagram in the bottom-left panel illustrates the possible clinical history of a particular patient, for each of waves EW-2 to EW-6, ranging from Day 1 to the last day the wave in question. The bottom-right panel presents the number of identified patients obtained from our patient tracking protocol in each combination of clinical stage and epidemiological wave.

All of our neural networks rely on a feed-forward architecture with two layers. The hidden and output layers comprise two sigmoid neurons and two softmax neurons, respectively. Cross-entropy was selected as the training cost [36–38], and the scaled conjugate gradient back-propagation method was used as optimizer [39, 40]. In our prospective study, we employ two distinct phases. In the first of these phases, we utilize the neural networks trained in Ref. [17], for each of the four clinical stages. We are then able to assess their performance when applied to patients in epidemiological waves occurring at dates later than those of the data used for training. Subsequently, in the second phase, we have trained neural networks using cumulative historical data covering the period up to the end of each of the waves in turn, and evaluated them at EW-2 through EW-5, irrespective of whether they lie within or outside the date span for the training data. To ensure a fair comparison in all cases, the neural network architecture is in all cases identical to that reported in Ref. [17].

## III. Results

As mentioned earlier, we have carried out our prospective study in two phases. As part of the first phase, we have conducted a patient-tracking protocol within each of the epidemiological waves. Specifically, we identify patients on day 1 of each of the waves who meet the criteria for stage 1 (see bottom panel of Fig. 2), namely those who seek medical evaluation and/or treatment and are suspected of harboring a COVID-19 infection, as yet unconfirmed either through a test or the appearance of COVID-19-related pneumonia symptoms. Note that this patient tracking procedure excludes any patients who may fall ill later than day 1 of each wave. These patients are then monitored day by day, until reaching the last day of the wave, taking note of any patients who may transition to stages 2,3, and 4. Note that patients in Stage 2 already have a defined COVID-19 status and may present COVID-19-related pneumonia symptoms, so the list of features is expanded accordingly. Those patients who transition to Stage 3 have been hospitalized, activating the hospitalization status feature. Patients in Stage 4 are those who unfortunaltey reach a critical state and have undergone intubation or have been admitted to an ICU unit, resulting in the activation of the corresponding features in their profiles.

The number of patients who start off at stage 1 in each wave (EW-2 through EW-5), as well as the number transitioning to each of stages 2, 3, and 4 are shown in the bottom panel of Fig. 2. Note that despite the severity of the pandemic, the number of patients transitioning to the higher stages is relatively low. Fortunately, the majority of patients do not suffer complications to be registered, and most remain at stage 1 until discharged. The fact that this analysis is restricted to those at stage 1 on day 1 of each wave, together with the low transition probability to higher stages, makes the number of patients reaching stage 4 quite small.

Note that our patient tracking protocol described above reflects the sequence of events that could be followed by a particular patient. Upon arrival of the patient at a clinic or hospital he or she is inspected at the triage, and from there transitions through some or all of the four clinical stages, with either recovery or death as the final outcome. It is at the triage stage that a predictive tool such as our neural networks becomes relevant: it can aid healthcare professionals to identify patients who are at higher risk, thus helping to more efficiently manage the hospital resources and capacity, and to provide timely treatment. In this regard, we utilized the neural networks trained in Ref. [17] to assess their predictive power using the patient data obtained during the tracking process. The resulting accuracy of these neural networks at each clinical stage and each epidemiological wave is recorded in Table III. The accuracy is calculated as the sum of true negatives and true positives divided by the total number of records.

**TABLE II.** Overall accuracy of the five sets of neural networks trained with cumulative historical data from each epidemiological wave at different clinical stages. These networks are tested during the second phase of the prospective study, see Figs. 3 and 4.

| Training Data | Stage |  |  |  |
| --- | --- | --- | --- | --- |
|  | 1 | 2 | 3 | 4 |
| Ref. [17] (January 31, 2021) | 84.3% | 90.5% | 93.1% | 93.5% |
| EW-2 (up to week 15 of 2021) | 84.8% | 91.0% | 94.2% | 94.2% |
| EW-3 (up to week 42 of 2021) | 85.1% | 91.6% | 95.0% | 95.1% |
| EW-4 (up to week 9 of 2022) | 87.7% | 91.6% | 95.0% | 95.1% |
| EW-5 (up to week 33 of 2022) | 85.1% | 91.6% | 96.3% | 96.4% |
| EW-6 (up to week 4 of 2023) | 86.1% | 91.5% | 96.3% | 95.5% |

**TABLE III.** Prospective overall accuracy of early-data-trained neural networks. The networks, described in Ref. [17], are tested with data obtained from the patient tracking process for each of the four clinical stages within every epidemiological wave.

|  | Stage |  |  |  |
| --- | --- | --- | --- | --- |
| Wave | 1 | 2 | 3 | 4 |
| EW-2 | 78.0% | 90.7% | 54.6% | 83.5% |
| EW-3 | 83.7% | 92.3% | 51.2% | 0% |
| EW-4 | 71.7% | 86.6% | 62.5% | 100% |
| EW-5 | 83.0% | 94.8% | 84.0% | 50% |
| EW-6 | 80.1% | 96.2% | 69.3% | N.A.% |

Because the networks from Ref. [17] were trained using data covering the period up to 31 January 2021, we have tested them on patients ranging from waves EW-2 to EW-6. The accuracies shown in Table III that correspond to Stages 1 and 2 present high efficiencies due to the large number of records in these stages. Stage 3 does not contain a sufficient number of records for the neural networks to detect patterns correctly, which leads to a markedly lower accuracy. This difficulty is compounded for stage 4, which exhibits a very small number of cases leading to poor statistics and the inability to correctly estimate the algorithm efficiency. The N.A. value appearing for stage 4 of EW-6 refers to the absence of records in this wave and stage.

In the second phase of our prospective study, we train five sets of neural networks using cumulative historical data covering the period up to the end of each wave (EW-2 through EW-5). We then evaluate their prediction effectiveness using data obtained from the patient tracking process. Through this study, we aim to ascertain the optimal number of waves to include in the training of neural networks for predicting high-risk COVID-19 patients with the highest accuracy possible.

We first describe how the second-phase neural networks are prepared. As was the case in the first phase of our study, we train a separate network for each of the four stages. The first set of four networks is trained with data covering the period up until the end of EW-2, i.e. week 15 of 2021. The second set of four networks is trained with data covering the period up to the end of EW-3, i.e. week 42 of 2021. This process has been repeated for data covering the periods up until the end of EW-4, EW-5, and EW-6, corresponding to week 9 of 2022, week 33 of 2022, and week 13 of 2023, respectively. As was done in Ref [17], 70% of the data was allocated for training, 15% for validation, and 15% for testing. The accuracy of each set of neural networks is shown in Table II.

To assess the predictive capabilities of our trained network set, we have tested it on the dataset obtained through the patient tracking protocol, for each wave, used in the first phase of our study. In figure 3 we plot the number of new confirmed patients plotted vs date throughout the date span covered by the database, as was also done in Fig, 1a. For each wave EW-2 through EW-5, and each of the four stages, we display the resulting accuracies of our networks in the gray boxes. In the five subpanels, (a) through (e), we employ for neural network training a progressively larger fraction of the total date span, colored in red (note that we have colored in blue the excluded data). Note that despite the enlargement of the training region, the accuracies remain remarkably consistent in all cases. More importantly, these accuracies are comparable to those achieved with the neural network proposed in Ref. [17]. These findings suggest that neural networks trained with historical data during the early stages of the pandemic exhibit strong predictive capabilities, which permits precise identification of high-risk patients in subsequent epidemiological waves.

**FIG. 3.**
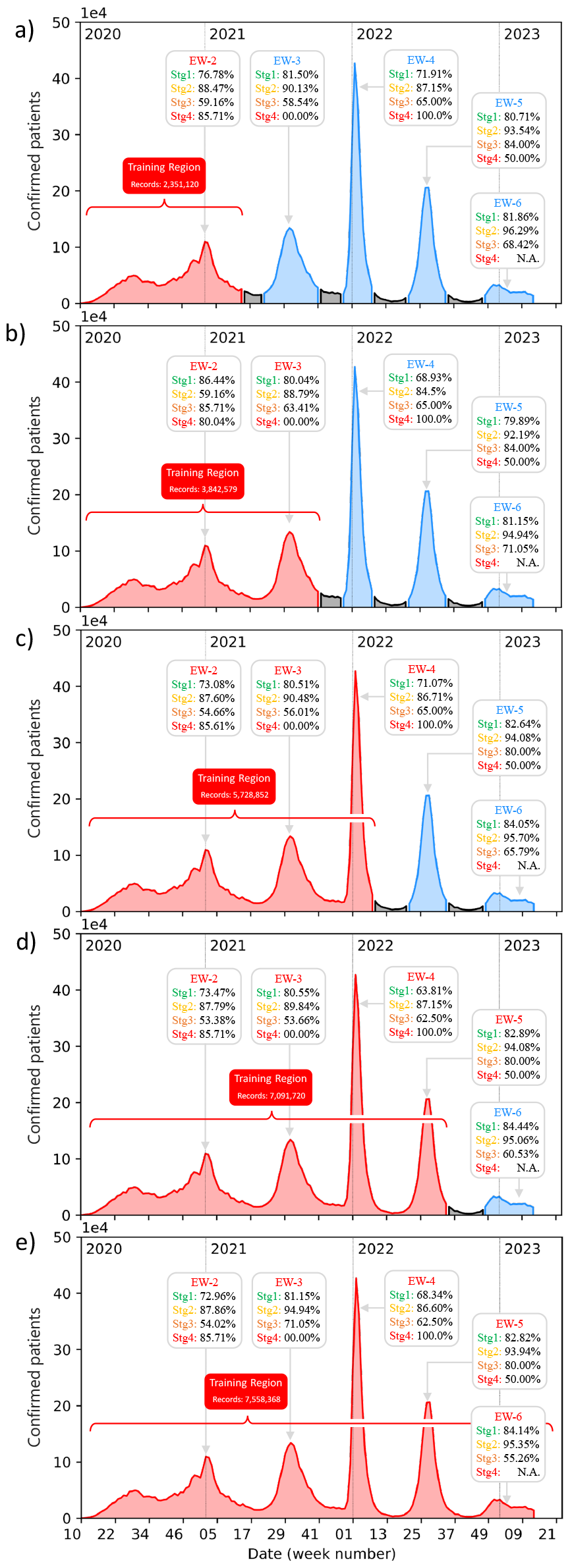
Prediction accuracy of our neural networks, calculated for patients in each of waves EW-2 through EW-6, for each of the four clinical stages. The length of the training period is progressively extended, adding one wave at a time, as displayed in the five subpanels so that in (e) data from all waves is used for training.

For the sake of completeness, and to explicitly show the robustness of early-data-trained neural networks, Figure 4 shows the accuracy of neural networks trained with increasingly larger datasets (including an increasing number of waves) when predicting patient clinical outcomes for patients in each of the stages, and in each of waves EW-2 through EW-6. Each subpanel, (a) to (e), shows the prediction accuracies of the neural network, with a structure identical to that used in Ref.[17], trained with data up to the epidemiological wave number indicated in the horizontal axis. The subpanel labels shown in the upper right corner denote the epidemiological waves to which the patient tracking protocol is applied to obtain the testing data. Note that Stage 4 (red line) of waves EW-3, 4, and 5 presents fluctuations, i.e. very high or very low prediction accuracies, which follows from the limited size of the respective dataset. Moreover, for Stage 4 of EW-6 we have no predictive value since no patient in the dataset reached this stage.

**FIG. 4.**
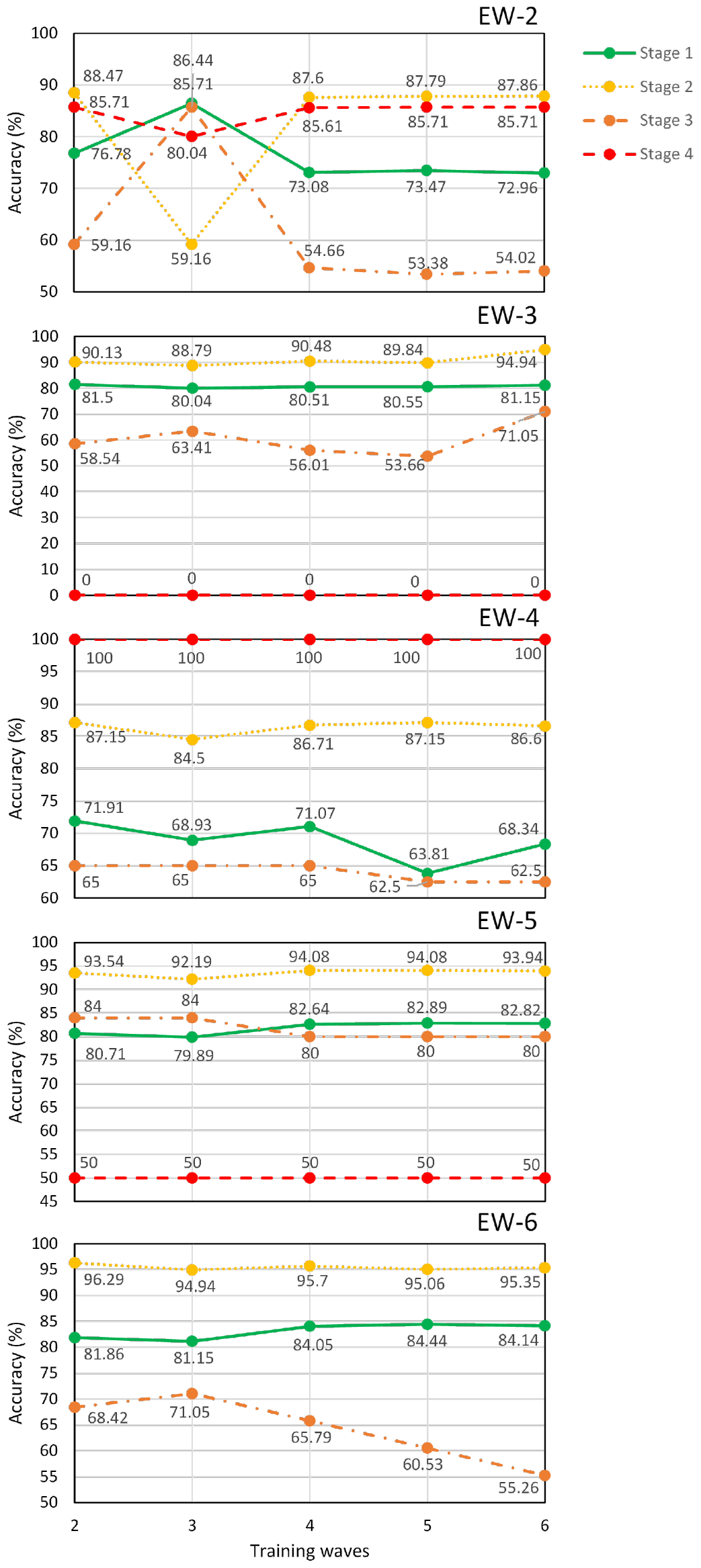
Prediction accuracy plotted, for each of stages 1-4 as labeled, vs the length of the training data date span, indicated in the horizontal axis as the most recent wave included. The accuracy values are obtained upon applying our neural networks to testing data resulting from our patient protocol tracking protocol; each subpanel indicates the use of data corresponding to each of waves EW-2 through EW-6.

Upon inspection of figure 4, it is apparent that (overall) the neural network efficiencies remain stable, for each of the stages, as one extends the date span of the training data. There are some notable exceptions, for example, in extending the date span from waves 2 to 4, for patients in our tracking protocol belonging to EW-2, an important fluctuation is observed. We believe that this may be related to the fact that the peak of EW-3 coincides with reaching the milestone of half of the population being vaccinated. Similarly, among patients in each of waves EW-2 through EW-6, those in stage 1 and EW-4 exhibit an abnormally low (compared to the other waves) prediction efficiency. We believe that this may be related to the appearance of the Omicron variant (with a higher infection rate but generally milder symptoms as compared to previous strains), which coincides with the onset of wave EW-4.

Finally, we remark that Figure 4 allows us to conclude that early-data-trained neural networks, such as the one used in Ref. [17], can predict with a precision greater than 84.5% the clinical outcome for patients who are tracked from Stage 2, i.e., patients with a confirmed diagnosis of COVID-19 who do not yet present respiratory complications. This may certainly allow for a more efficient allocation of resources for such patients. We believe that the results of this prospective study provide the building blocks for novel strategies to predict outcomes in clinical settings. Moreover, they provide new tools to be exploited for clinical decision-making in the context of future large-scale epidemic or pandemic events.

## IV. Conclusions

Although the COVID-19 crisis has by now taken a backseat in current world affairs, it is of utmost importance to provide health-sector professionals with technological tools that allow them to effectively manage available clinical resources during possible future pandemics or large-scale epidemic events. In Ref. [17] we proposed state-of-the-art machine-learning models that can, with high efficiency, identify high-risk COVID-19 patients across four clearly-identified clinical stages. In this work, we have presented a prospective validation study, based on tracking individual patients day-to-day in each of the latter five COVID-19 epidemiological waves, among the six officially recognized waves in Mexico. We apply our machine-learning models from Ref. [17] to the patients in datasets obtained from our patient tracking protocol. On the one hand, we employ the same networks reported in [17] (without any retraining), and apply them prospectively to patients in each of waves EW-2 through EW-6. On the other hand, we retrain the networks with data covering an increasing date span to include a successively larger number of epidemiological waves and apply prospectively these re-trained networks to patients in each of waves EW-2 through EW-6. Our results show that models trained with early historical data demonstrate significant predictive capabilities permitting precise identification of high-risk patients in subsequent epidemiological waves, with efficiencies in line with those obtained with networks trained with data from an extended date span. We are certain that these results establish the grounds for innovative strategies in predicting individual clinical outcomes in the context of epidemiology, providing valuable insights for possible future health crises.

## Acknowledgments

M.A.Q.-J. thankfully acknowledges financial support by CONAHCyT under the Project CF-2023-I-1496 and by DGAPA-UNAM under the Project UNAM-PAPIIT TA101023. R.J.L.-M. thankfully acknowledges financial support by DGAPA-UNAM under the project UNAM-PAPIIT IN101623. A.B.U. thankfully acknowledges financial support by DGAPA-UNAM under the project UNAM-PAPIIT IN103521.

## Data Availability

The data that support the findings of this study are openly available in the Statistical Morbidity Yearbooks published by the General Council of Epidemiology, part of the Health Ministry, Mexican Federal Government. Data are available in https://www.gob.mx/salud/documentos/datos-abiertos-bases-historicasdireccion-general-de-epidemiologia.

